# Proteomic signatures of perioperative oxygen delivery in skin after major surgery

**DOI:** 10.1101/2020.08.25.20181859

**Authors:** Gareth L. Ackland, Emily Bliss, Fatima Bahelil, Trinda Cyrus, Marilena Crescente, Timothy Jones, Sadaf Iqbal, Laura Gallego Paredes, Andrew J. Toner, Ana Gutierrez del Arroyo, Wendy E. Heywood, Edel A. O’Toole, Kevin Mills

**Author notes:** Correspondence to: G L Ackland, Reader in Perioperative Medicine, Translational Medicine & Therapeutics, William Harvey Research Institute, Queen Mary University of London, EC1M 6BQ. United Kingdom.

## Abstract

**Objective:** Maintaining adequate oxygen delivery after major surgery is associated with minimizing organ dysfunction, although the precise molecular mechanisms remain unclear.

**Background:** Skin, the largest organ in the body, is vulnerable to reduced oxygen delivery. We hypothesized that the skin proteome, assessed before and after surgery, would reveal molecular differences in patients randomized to receive cardiovascular therapy aimed at maintaining preoperative oxygen delivery (DO_2_).

**Methods:** Abdominal punch skin biopsies were snap frozen or fixed in paraformaldehyde immediately before and 48h after elective esophageal or liver resection. Immediately after surgery, patients were randomized to standard of care or therapy to maintain preoperative DO_2_. On-line two-dimensional liquid chromatography, followed by ultra-high definition label-free mass spectrometry analysis, and/or immunoblots quantified significant proteomic changes. Selected proteins identified by mass spectrometry were confirmed by immunohistochemistry and immunoblot. In a murine hepatic resection model, confirmation of specific proteomic signatures identified in patients was sought by immunoblotting.

**Results:** Paired biopsies were analyzed from 35 patients (mean age:68±9y; 31% female). We identified 2096 proteins, of which 157 were differentially expressed after surgery. Similar results for selected proteins were found using immunohistochemistry (n=6 patients), immunoblots (n=12 patients) and murine abdominal skin obtained after liver resection (n=14). After surgery, 14 proteins distinguished esophagectomy patients with normal (n=10) versus low (n=7) DO_2_.values. Failure to maintain preoperative DO_2_ was associated with upregulation of proteins counteracting oxidative stress and.

**Conclusions:** Serial skin biopsies afford mechanistic insight into end-organ injury by quantifying proteomic changes associated with impaired oxygen delivery during high-risk surgery.

**Trial registration:** ISRCTN76894700

**Funding:** Academy of Medical Sciences/Health Foundation Clinician Scientist Award [GLA]; British Oxygen Company research chair grant in Anesthesia [GLA]; Great Ormond Street Hospital Biomedical Research Centre [EB,WH,KM]; British Heart Foundation:PG/17/40/33028 [MC]; UK NIHR [GLA]; Barts Charity [TJ]

**Mini-Abstract:** Skin, one of the largest organs in the body, is vulnerable to reduced oxygen delivery. Proteomic analysis of skin biopsies obtained before and after surgery show distinct metabolic and inflammatory changes related to perioperative oxygen delivery. Mirrored by complementary laboratory data in mice, skin proteomics reveal new insights into perioperative organ dysfunction.

## Introduction

Major complications after elective esophagectomy and other major surgeries including liver resection occur in up to ~20% of patients, resulting in delayed discharge and/or readmission to hospital. ^1,2^ The development of complications after major surgery is a key determinant of survival.^3^ Maintaining oxygen delivery is strongly associated with minimising organ dysfunction and reducing morbidity after surgery.^4-7^ The intersection between metabolic dysfunction and inflammation may underpin this link,^8,9^ but the molecular mechanisms remain unclear. In part, advancing mechanistic insight into perioperative organ injury is hindered by the lack of serially obtained, readily accessible tissue samples from end-organs.^10^

Preserving blood flow is necessary for preserving organ function, including skin integrity and wound healing after surgery.^11^ Skin is particularly vulnerable to reductions in cardiac output and hence oxygen delivery, as blood is preferentially shunted away from skin to preserve the perfusion of vital organs.^12^ Direct injury to the skin triggers an immune response that coordinates wound healing.^13^ However, even after distant injury, cytokine production is up-regulated in uninjured skin from a range of skin-resident cells.^14^ Thus, skin-derived mediators are likely to contribute to the postoperative modulation of systemic immunity and organ dysfunction through lymph node drainage.^15^

Although readily accessible, the high lipid content, insolubility and extensive cross-linking of proteins in skin has presented major challenges for proteomic analysis. However, recent advances in skin proteomics have overcome these technical challenges, thereby enabling previously unattainable mechanistic insights into various dermatologic pathologies.^16^ To further explore molecular mechanisms underlying end-organ injury after surgery, we therefore examined the relationship between skin proteomics and oxygen delivery. Serial skin punch biopsies were obtained from high-risk surgical patients in a randomised controlled trial which demonstrated that morbidity was less frequent in patients in whom preoperative oxygen delivery values was maintained after surgery (regardless of goal-directed haemodynamic therapy).^6^ As well as characterising general proteomic changes 48h after surgery, this approach added mechanistic insight into the impact of early hemodynamic management on end-organ cellular processes following major noncardiac surgery.

## Methods

### Patient participants

POM-O (Postoperative Morbidity Oxygen delivery) was a multicentre, double-blind, randomised controlled trial^6^ done in four university hospitals in the UK (ISRCTN76894700) and was approved by the South London Research Ethics Committee Office (09/H0805/58). Eligibility criteria have been published in the original trial report and are provided in Supplementary data. Adult patients undergoing major elective surgery gave written informed consent for skin biopsy samples to be obtained. Patients were allocated by computer-generated randomisation to a postoperative protocol (intravenous fluid, with and without dobutamine) targeted to achieve their individual preoperative oxygen delivery value (goaldirected therapy) or standardised care (control). Patients and staff were masked to the intervention. The primary outcome was absolute risk reduction in morbidity (defined by Clavien-Dindo grade II or more)^17^ on postoperative day 2.

### Mouse model

To establish the translational potential for findings from patient skin biopsies, we examined proteomic changes by immunoblot in ventral abdominal skin obtained from male mice 48 hours after hepatectomy under isoflurane general anesthesia. C57BL/6 male mice aged 10–12 weeks (Charles River Laboratories) were maintained in accordance with the UK Home Office Legislation (1986 Animal Procedures Act). All procedures were approved by the Institutional Animal Care and Use Committee at William Harvey Research Institute. Experiments were performed and reported in accord with ARRIVE guidelines. Mice acclimatised for at least 48 hours prior to experiments, with access to food and water ad libitum.

Mice were randomised to receive specified interventions, using Research Randomizer. We compared skin samples from mice that underwent hepatectomy with naïve (non-operative) mice and mice that also underwent hair removal under general anesthesia over the same timeframe. In sterile conditions, mice were placed on a warming blanket. The abdomen was sterilised with chlorhexidine after hair removal cream was applied, before a midline abdominal incision and 50% hepatectomy was performed.^18^ Continuous vicryl suture was used to close the abdominal muscle, and interrupted prolene sutures were used to close the skin. Subcutaneous 0.9% saline (1ml) was administered at the end of the procedure. Chirocaine local anesthetic was injected around the incision site to ensure analgesia before mice recovered in a warming box until mobile. Forty-eight hours later, the mice were killed by cervical dislocation and abdominal skin removed.

### Skin biopsy and sample preparation

2mm skin punch biopsies (Instrapac, Robinson Healthcare, Worksop, UK) were obtained from surgical patients within 5 cm of the surgical incision within 5 seconds of inducing general anesthesia, and 48 hours after surgery at a similar location on the opposite side of the surgical incision (under local anesthesia). Punch biopsies were immediately snap frozen in liquid nitrogen or immersed in 4% paraformaldehyde. For proteomic analyses, samples were prepared as described previously.^16^ Briefly, snap-frozen samples were homogenised in 50 mM ammonium bicarbonate and 2% w/v ASB-14 using bead (Precellys 1.4mm diameter ceramic beads, Peqlab, VWR) and mechanical homogenisation (Minilys®, Bertin Technologies). A modified Lowry protein assay (Pierce™, ThermoFisher Scientific) was used to calculate the protein content and 50 μg was lyophilised using a freeze drier. Proteins were resolubilised in 100 mmol/L TRIS-HCL, pH 7.8, containing 2 % w/v ASB-14, 6 mol/L urea and 2 mol/L thoiurea and digested using Sequencing Grade Modified Trypsin (Promega, Madison, USA).

### Two-dimensional liquid chromatography

Following in-solution digestion of the homogenised sample, purification of the digested sample peptides reverse phase C-18 chromatography was undertaken to remove the salt and lipid content of the sample (ISOLUTE® C18 columns, Biotage), as described previously.^16^ Then, two dimensional high pH fractionation of sample peptides, followed by low pH chromatographic separation using an online nanoAcquity ultra high performance liquid chromatography system was undertaken (Supplementary data). For each fraction, a 60 min mass spectrometry analysis was performed on a SYNAPT G2-Si (Waters, Manchester, UK) mass spectrometer (Waters, Manchester, UK) in a UDMS^E^ mode in positive ion electrospray ionisation mode.^19^

### Proteomic analysis

Raw mass spectrometry data were processed using Progenesis QI analysis software (Nonlinear Dynamics, U.K.). Peptide identifications were performed using MS^e^ search identification against the Uniprot Human reference proteome 2015, with 1 missed cleavage and 1% peptide false discovery rate (FDR).^20^ Fixed modifications were set to carbamidomethylation of cysteines and dynamic modifications of hydroxylation of aspartic acid, lysine, asparagine and proline and oxidation of methionine. A protein was considered to be differentially expressed if there was a significant change (p<0.05) from preoperative values accounting for the degree, direction, and rank of difference between samples obtained before and 48h after surgery with sequence length ≥6, hits ≥ 2, a maximal mean fold change ≥1.5 and >20-confidence score, a further measure of false-discovery rate (Progenesis QI). Proteomics data was deposited in the PRIDE PRoteomics IDEntifications (PRIDE) public-domain repository, which provides a single point for submitting mass spectrometry based proteomics data.^21^ Data was normalised and significance determined by one-way ANOVA analysis (Progenesis QI software).

### Immunohistochemistry

Immunofluorescence on frozen sections of human skin biopsies was used to confirm selected findings from the proteomic data by histologists masked to clinical details (Atlantic Bone Screen, Cedex, France). Using validated antibodies (Supplementary data), protein expression of integrin alpha-6 (ITGA6; 1:50; Merck Millipore (MAB1378)) and the cell surface glycoprotein CD44 (1:50; Novocastra (BMS144)) were quantified by immunofluorescence by an investigator masked to clinical details (Fiji software, NIH Image, USA).

### Immunoblots

Membranes were incubated with the following species-specific primary antibodies for parkin and superoxide dismutase-1 to confirm selected findings from the proteomic data, which were masked to clinical details. Secondary antibodies (1:2000; Dako, Stockport, UK or Cell Signaling Technology, Leiden) were rabbit anti mouse HRP or goat anti-rabbit HRP, as indicated. Membranes were developed using ECL^TM^ reagent and Hyperfilm ECL (Amersham, UK).

### Statistical analyses

All analyses were conducted by investigators blinded to the identity of treatments/genotypes. Clinical data were processed by analysers blinded to patient group assignments. For all statistical tests, the Benjamin-Hochberg procedure was applied where applicable to correct for testing multiple hypotheses. Data distribution was assessed by the Kolmogorov-Smirnov normality test. P values < 0.05 were considered significant.

### Sample size estimation

Sixteen paired patient samples were estimated to be required to have a 90% chance of detecting a 42±25 difference in the number of proteins with ≥1.5-fold change detected before versus after surgery (confidence score>20; α=0.05; 1-β=0.9; Sealed Envelope).

### Bioinformatic analyses

We assessed categorical changes in skin protein expression (as defined by fold-change and confidence scores) before and after surgery using the STRING database, which collects, score and integrates all publicly available sources of protein-protein interaction information through which computational predictions for direct (physical) as well as indirect (functional) interactions are made.^22^ For the enrichment analysis, STRING employs the established classification systems Gene Ontology ^23^ and KEGG^24^ but also offers additional, new classification systems based on high-throughput text-mining as well as on a hierarchical clustering of the association network itself. To compare quantitative fold-changes in protein expression between oxygen delivery achievers versus non-achievers, we used Ingenuity Pathways Analysis (IPA) (01-13) software (QIAGEN, USA) to perform enrichment analyses^25^ to estimate the significance of observing a candidate protein set within the context of biological systems.^26^ Briefly, protein identifiers were mapped in the Ingenuity Pathway Knowledge Base (IPKB) to find cellular functions and diseases significantly associated with differentially expressed proteins. We determined changes in biological processes associated with differential protein expression by using downstream effect analysis. Molecular interactions between proteins and affected functions were visualised using network analysis.

## Results

### Patient characteristics

Punch skin biopsy samples were obtained from 35 patients who underwent esophagectomy or liver resection (mean age:68±9y; 31% female) before and 48h after surgery (Table 1). No biopsy sites became infected. Label free proteomic analysis was undertaken in 17 patients who underwent esophagectomy, of whom 7 failed to achieve their preoperative DO_2_ target (Figure 1). 7/8 (87.5%) patients who failed to achieve their preoperative DO_2_ target were more likely to sustain serious complications (including sepsis) after surgery, compared to 14/27 (51.8%) patients who achieved their individualised preoperative DO_2_ target (absolute risk reduction:36% (95% confidence intervals:6 to 65%)).

**Table 1:** Patient characteristics. Data is presented as mean with standard deviations (SD) for parametric data and as median (25th-75th interquartile range) for non-parametric data. Frequencies are presented with percentages (%). Age is rounded to the nearest year. *Indication for surgery was for treatment of cancer. Statistical analysis using paired-sample t-test or Wilcoxon-Signed rank test for continuous data and Chi-Squared test for categorical data.

|  | Achiever<br>n=27 | Non-achiever<br>n=8 |
| --- | --- | --- |
| Age (yr) | 70 (61-77) | 67 (65-70) |
| Female sex (n;%) | 4 (31%) | 3 (43%) |
| Body Mass Index (kg m<sup>-2</sup>) | 27.2 (4.3) | 28.8 (2.8) |
| Hemoglobin (g dL<sup>-1</sup>) | 124 (18) | 140 (18) |
| Albumin (g dL<sup>-1</sup>) | 42 (5) | 45 (2) |
| **Co-morbidities (n; %)** |  |  |
| Cardiovascular disease | 22 (81%) | 5 (63%) |
| Diabetes Mellitus | 7 (26%) | 0 |
| Hypertension | 12 (44%) | 3 (38%) |
| Malignancy* | 21 (78%) | 7 (88%) |
| **Intraoperative** |  |  |
| General + epidural anesthesia | 22 (82%) | 8 (100%) |
| Duration of operation (mins) | 300 (210-366) | 309 (290-374) |
| Fluid therapy (ml.kg.h) | 12.2 (6.8) | 9.1 (1.6) |
| Packed red cells (n;%) | 5 (19%) | 2 (25%) |
| Postoperative lactate (mmol L<sup>-1</sup>) | 2.3 (1.2-3.0) | 1.9 (1.7-2.4) |
| Postoperative Hemoglobin (g dL<sup>-1</sup>) | 110 (19) | 117 (22) |
| Oxygen delivery (% preoperative value) | 117 (100-159) | 77 (65-84) |
| Length of stay (days) | 17 (12-28) | 38 (17-91) |

**Figure 1.**
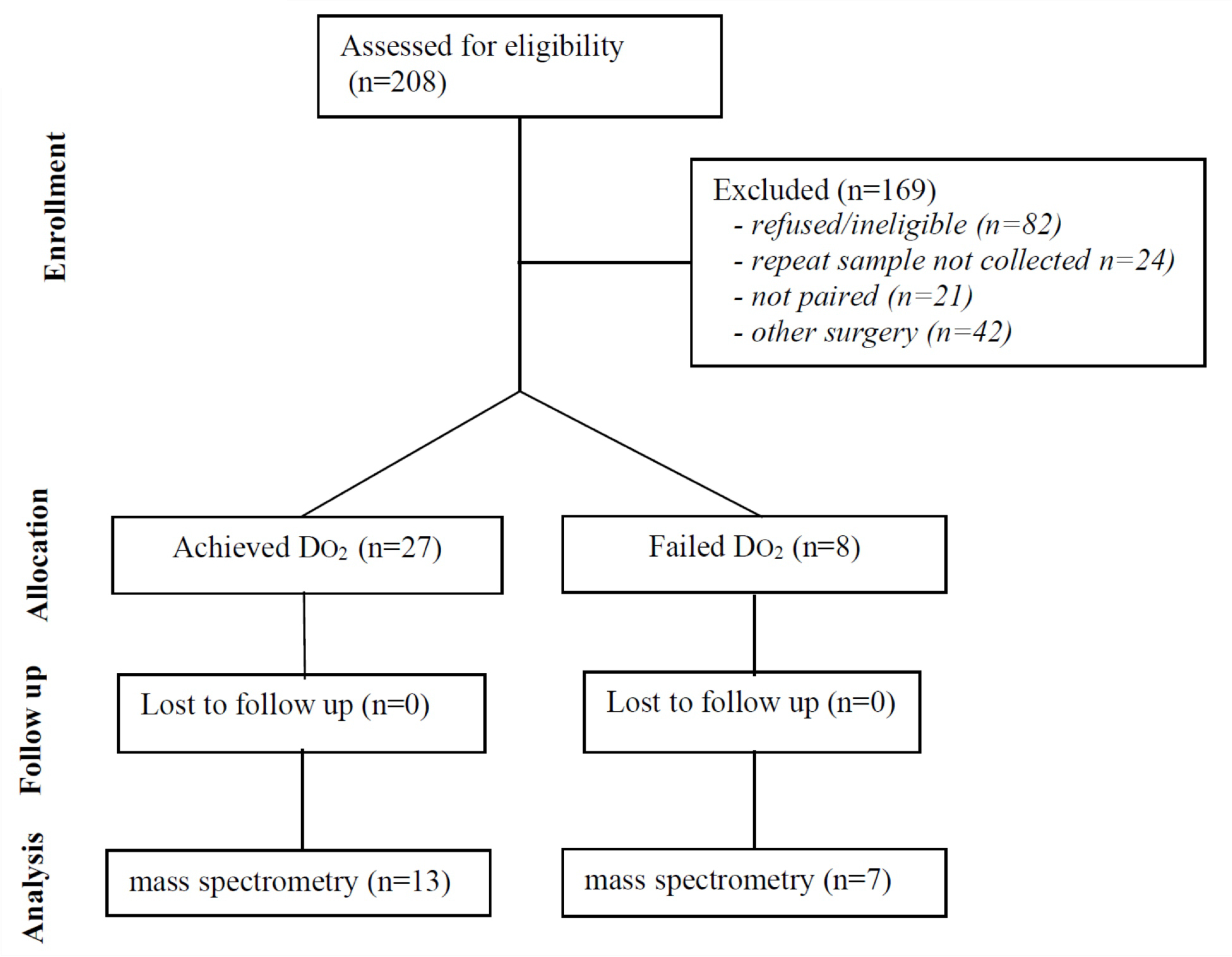
Flow diagram showing patients undergoing esophagectomy or liver resection eligible for sample acquisition who participated in the POM-O randomized controlled trial.

### Proteomic changes in skin after major surgery

We identified 2096 proteins in skin biopsy samples, of which 157 were differentially expressed (p<0.05) by at least 1.5-fold after surgery (median confidence score:124 (62-252); Figure 2A). Expression of 42 proteins increased after surgery (median fold change:2.31 (1.74-3.63)), whereas expression of 115 proteins declined after surgery (median fold change:1.89 (1.68-2.32); Figure 2B). Mirroring plasma values, in skin biopsy samples C-reactive protein increased (p=0.037; Figure 2B), whereas albumin decreased, after surgery (p=0.034; Figure 2C). After surgery, bioinformatic analyses revealed functional enrichment for major histocompatibility complex I antigen presentation (Reactome; p=0.034) and disruption of intermediate filament protein assembly (Gene Ontology; Figure 2D). A complete list of upregulated and downregulated proteins is provided in the supplementary data.

**Figure 2.**
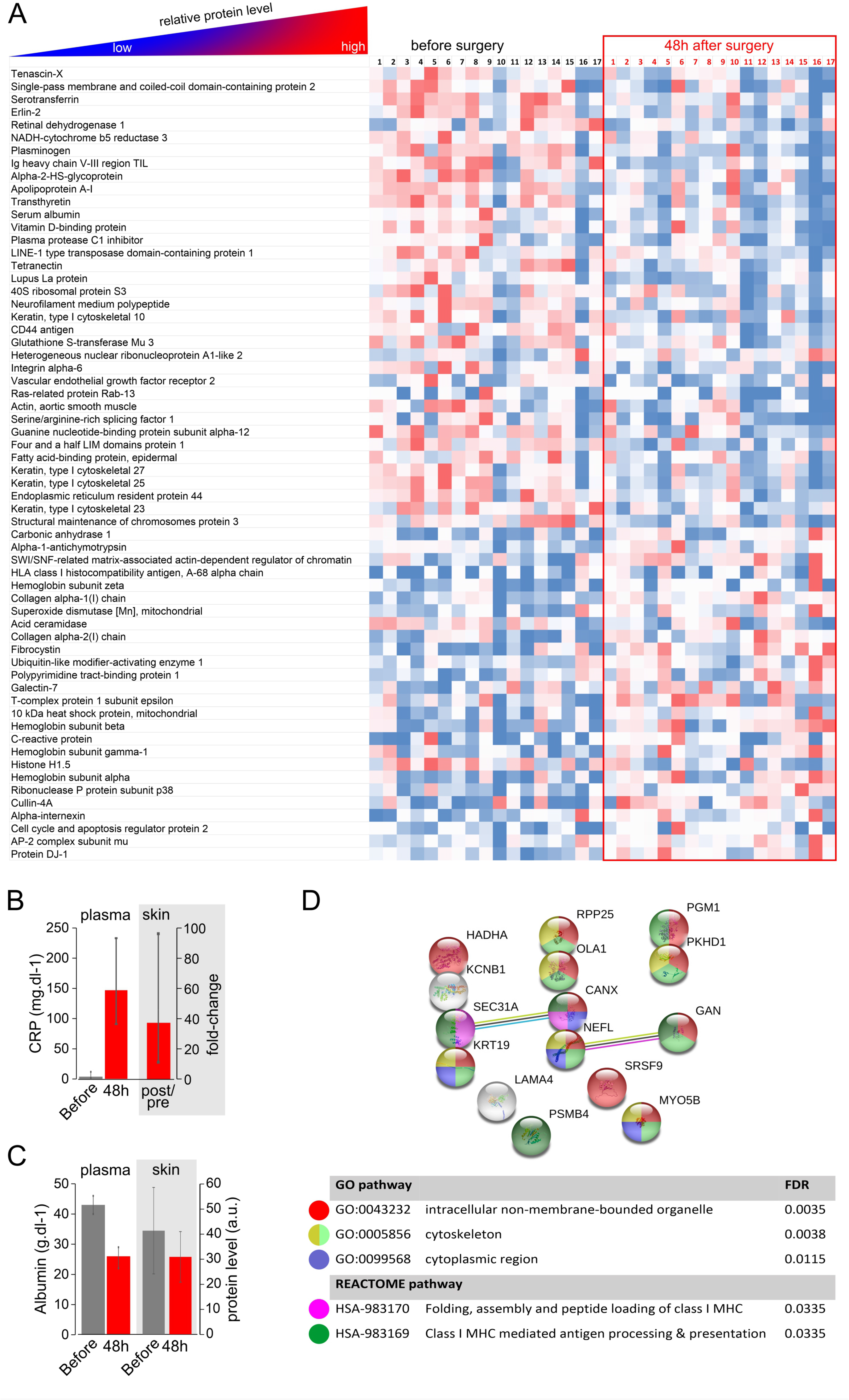
Differential expression of proteins in skin biopsies detected by mass spectrometry after major surgery. 68 proteins were differentially expressed by at least 1.5-fold (median confidence score:124 (62-252). Expression of 22 proteins increased after surgery (median fold change:2.27 (1.65-3.07)), whereas expression of 41 proteins declined after surgery (median fold change:1.75 (1.77-1.98)). Heatmap data is shown for relative changes for within protein comparisons. Mirroring plasma values, C-reactive protein also increased in skin biopsy samples (p=0.037, by paired t-test; n=17 patients). Albumin decreased in plasma and skin biopsy samples after surgery (p=0.034, by paired t-test; n=17 patients). After surgery, bioinformatic analyses revealed functional enrichment for MHC Class I antigen presentation (Reactome; p=0.034) and disruption of intermediate filament protein assembly (Gene Ontology; Figure 2E). Colour codes for each protein represent GO cellular site of action and/or REACTOME defined processes.

### Immunohistochemical and immunoblot validation of proteomic findings

To validate the findings of the derivation cohort of skin biopsy samples analyzed by mass spectrometry, we randomly selected skin biopsies from six individuals obtained before and after surgery for protein quantification by immunohistochemistry. We focused on proteins identified by mass spectrometry with established antibodies that have been validated for skin immunohistochemistry (Figure 3A). Expression levels of CD44, the principal cell surface receptor for hyaluronate, and α6β4 integrin, which stabilizes skin through the formation of hemidesmosomes, were reduced after surgery (Figure 3B). These changes were mirrored by semi-quantitative analysis by immunoblot (Supplementary data).

**Figure 3.**
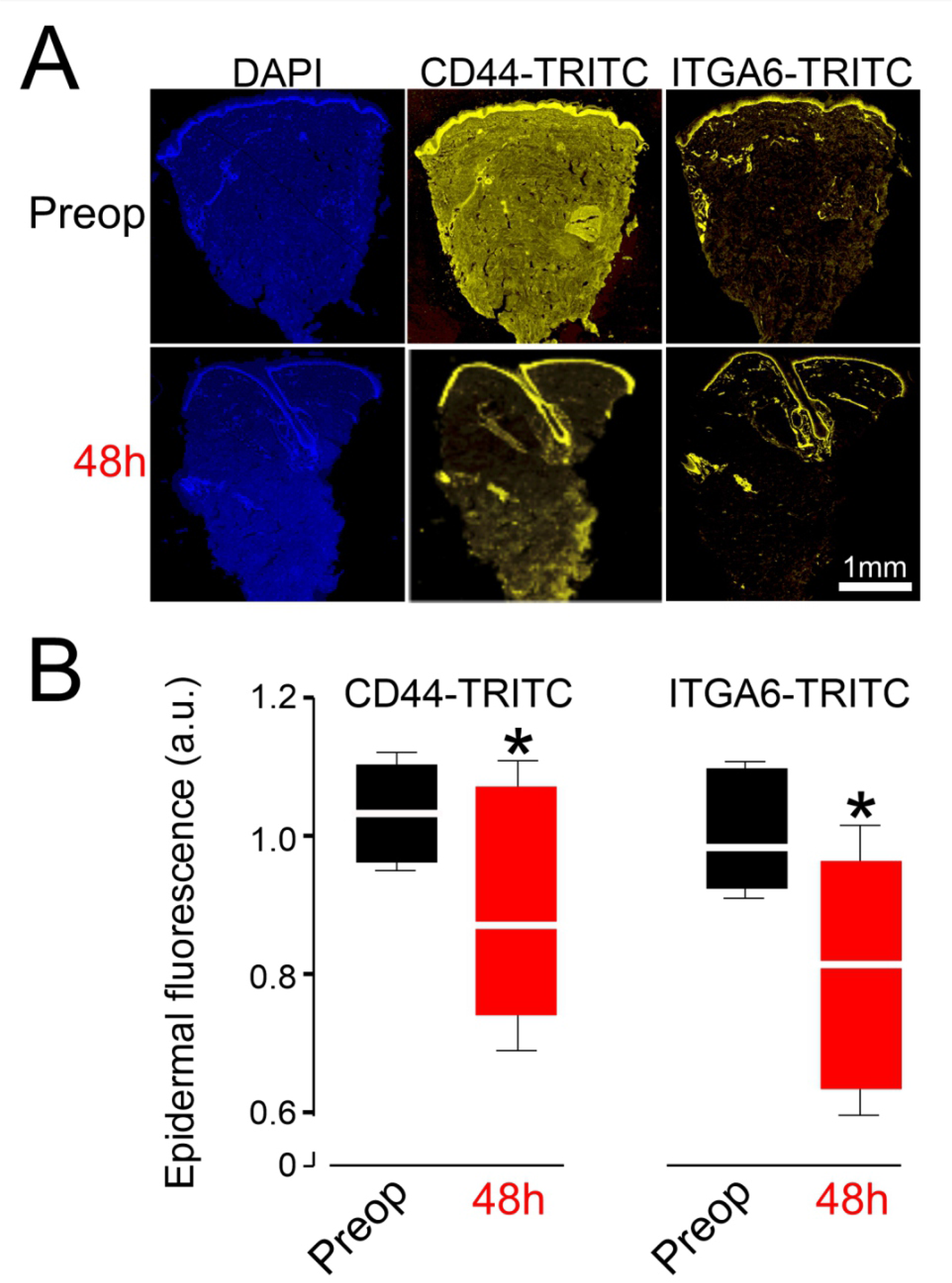
Immunohistochemistry validation of selected proteins. A. TRITC (tetramethylrhodamine) fluorescence for CD44 and ITGA6 was compared before and 48h after surgery in skin samples added to 10% formalin immediately after punch biopsy was performed. DAPI (4′,6-diamidino-2-phenylindole) staining identifies cell nuclei. Scale bar:100µm B. Summary data for epidermal fluorescence for CD44 and ITGA6 proteins, compared before and after surgery (standardised to group mean fluorescence levels before surgery; n=6 patients).

### Skin protein changes in murine surgical model

We next ascertained whether the proteomic changes after major surgery are also present in a an established translational murine model of higher-risk surgery- 50% hepatectomy (Figure 4A). We performed immunoblots of abdominal skin proteins using selected validated murine antibodies that we had identified in surgical patients by proteomics. After hepatectomy (n=8 mice), we observed that reduced expression levels of parkin (Figure 4B), the S-nitrosylation of which is regulated by DJ-1,^27^ were accompanied by higher expression levels of superoxide mutase-1 (Figure 4C). These data reflected the same pattern as that observed using mass spectrometry in surgical patients. The expression of these proteins was similar between naïve mice and those anesthetized who underwent abdominal hair removal, so skin proteomic changes were predominantly affected by the physiological changes accompanying hepatic resection rather than the local application of potentially inflammatory hair removal cream.

**Figure 4.**
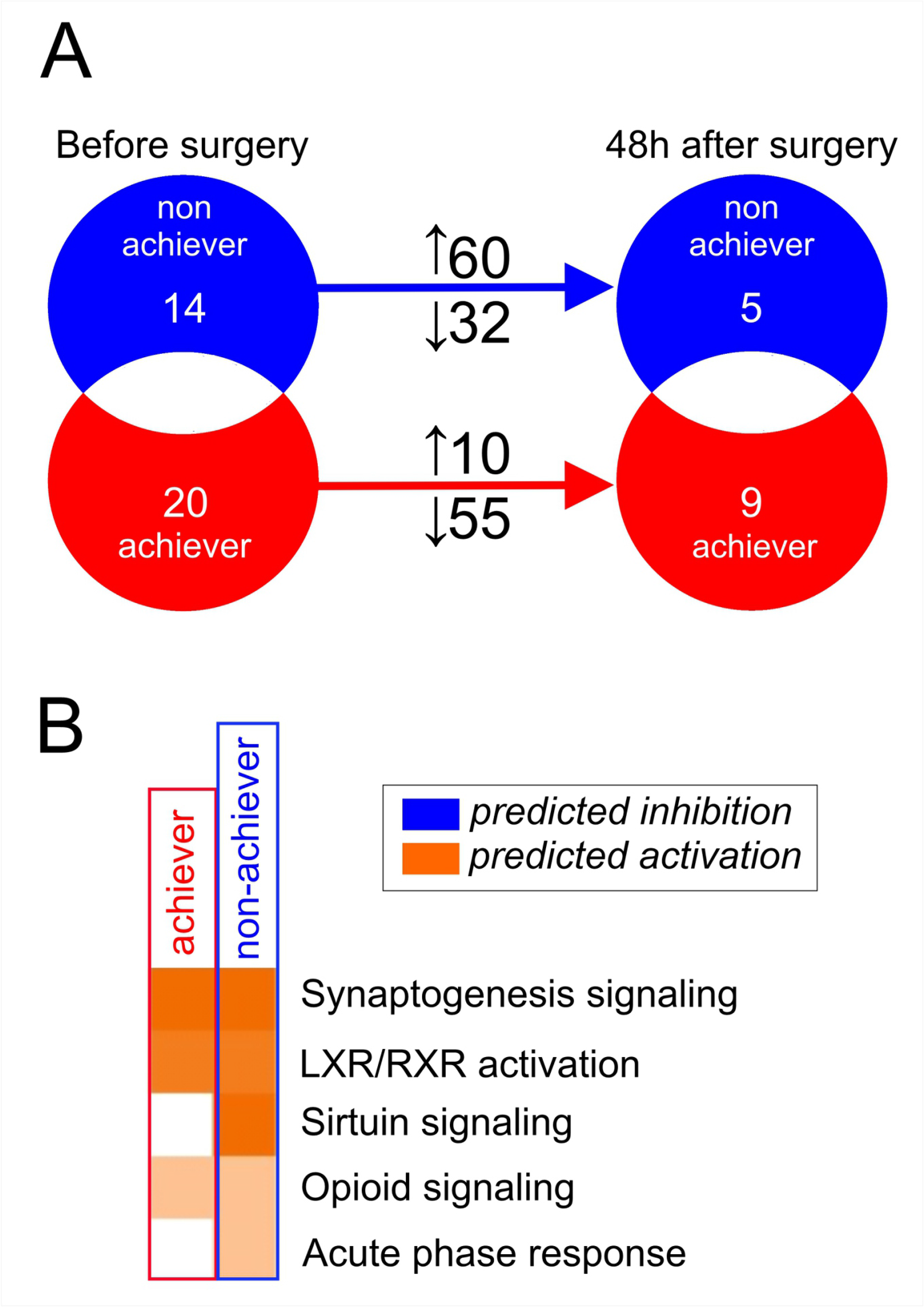
Selected skin proteome changes in murine model of 50% hepatectomy. **A.** 50% liver resection was undertaken in C57/Bl6 mice (n=8). Naïve mice and mice anesthetized for abdominal hair removal were compared with mice that underwent hepatectomy, to rule out non-specific inflammatory skin proteomic changes associated with the local application of hair removal cream (n=6). **B**. Immunoblots for parkin and superoxide dismutase-1, using beta-actin as loading control. Molecular weight markers are shown on right of blot. Original uncropped blot provided in supplementary data. **C**. Population data summarising immunoblot data for parkin after surgery. **D**. Population data summarising immunoblot data for superoxide dismutase-1 after surgery. Not that for both graphs, naïve mice and mice that underwent hair removal under isoflurane anesthesia are grouped together as controls, as there were no differences between them.

### Proteomic differences in skin associated with perioperative oxygen delivery

In patients where preoperative DO_2_ was achieved (n=10), the expression of 65 proteins differed after surgery (Figure 5A). The median expression of 55 proteins was reduced 1.89fold (1.62-2.23), compared with 10 proteins in which expression increased 1.92-fold (1.64-2.51) after surgery. In patients who failed to achieve preoperative DO_2_ values (DO_2_ non-achievers; n=7), 92 proteins changed after surgery (median confidence score:116 (62-223); supplementary data). The median expression of 60 proteins increased 1.9-fold (1.7-2.3), with 32 proteins decreasing after surgery by 2.54-fold (1.81-4.83). Heatmaps illustrating the proteins identified in each patient group are shown in supplementary data. Canonical pathways for acute phase response and sirtuin signaling proteins were enriched in non-achievers, compared to achievers (Figure 5B).

**Figure 5.**
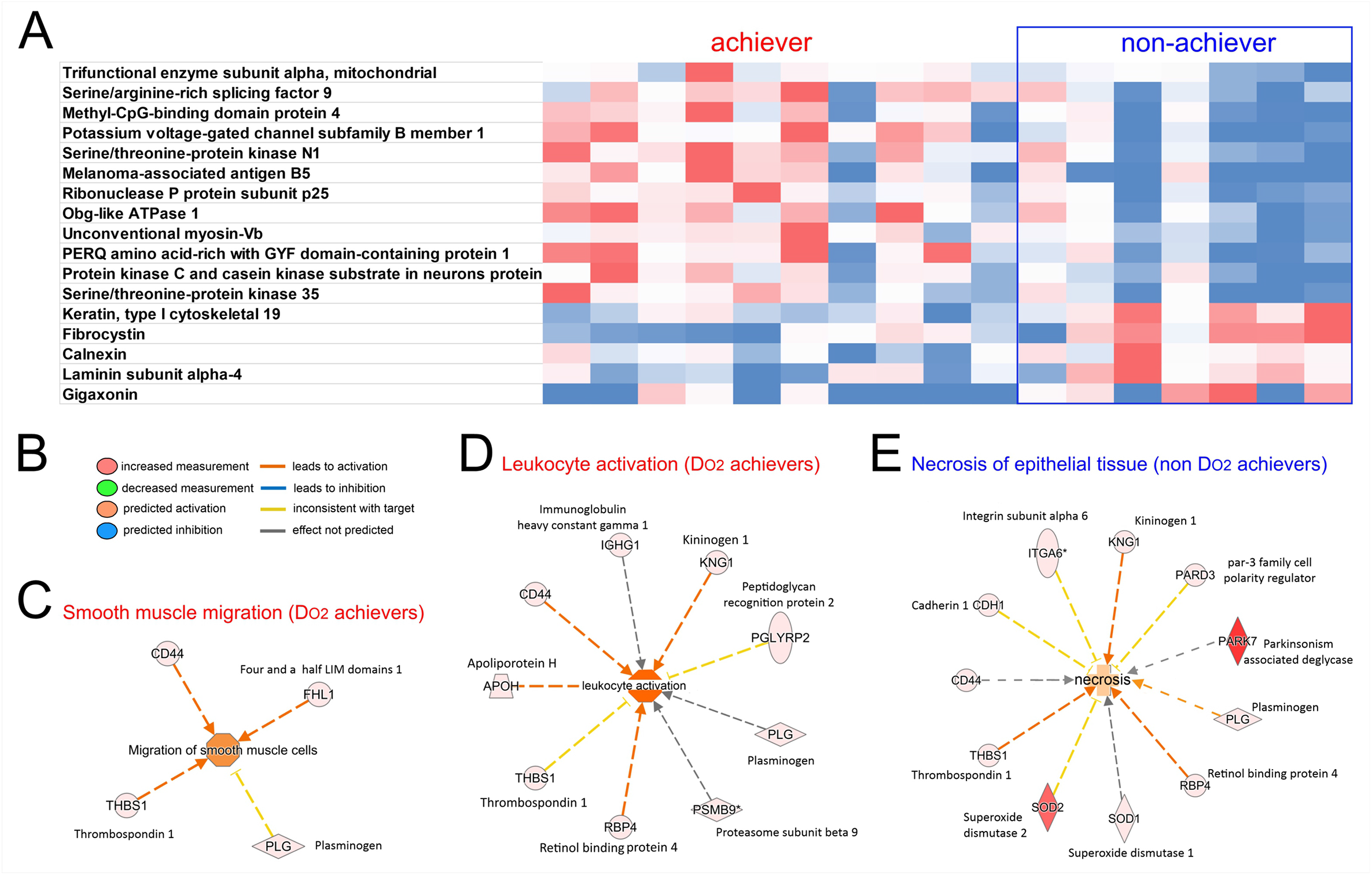
Proteomic differences in skin associated with perioperative oxygen delivery. A. Venn diagram showing directional changes in protein expression for patients who achieved (n=10), or failed to achieve (n=7) their individualized preoperative DO_2_ value. An increase/decrease was defined by fold-change >1.5, confidence score >20 and p value <0.05 (ANOVA), as generated by Progenesis QI analysis software. B. Canonical pathway analysis for enriched signaling categories for patients who achieved, or failed to achieve their individualized preoperative DO_2._ value.

### Systems biology analysis

After surgery, 15 proteins were differentially expressed between non- DO_2_ achievers and DO_2_ achievers (Figure 6A; median confidence score:63.7 (27.1-128.5)). To further dissect potential molecular interactions using Ingenuity Pathway Analysis (Figure 6B), we directly compared the 157 proteins identified by significant fold-changes after surgery for both DO_2_ achievers and non- DO_2_ achievers. This bioinformatic approach revealed that proteins involved in smooth muscle migration (Figure 6C) and leukocyte activation (Figure 6D) pathways were upregulated in patients where the perioperative oxygen delivery target was achieved. By contrast, in patients who failed to achieve their preoperative DO_2_, proteins involved in epithelial necrosis (Figure 6E) with established roles in oxidative stress (superoxide dismutase-1, DJ-1/PARK7) and wound healing (CD44, thrombospondin, plasminogen, retinal binding protein-4) were differentially expressed compared to achievers (Supplementary data).

**Figure 6.**
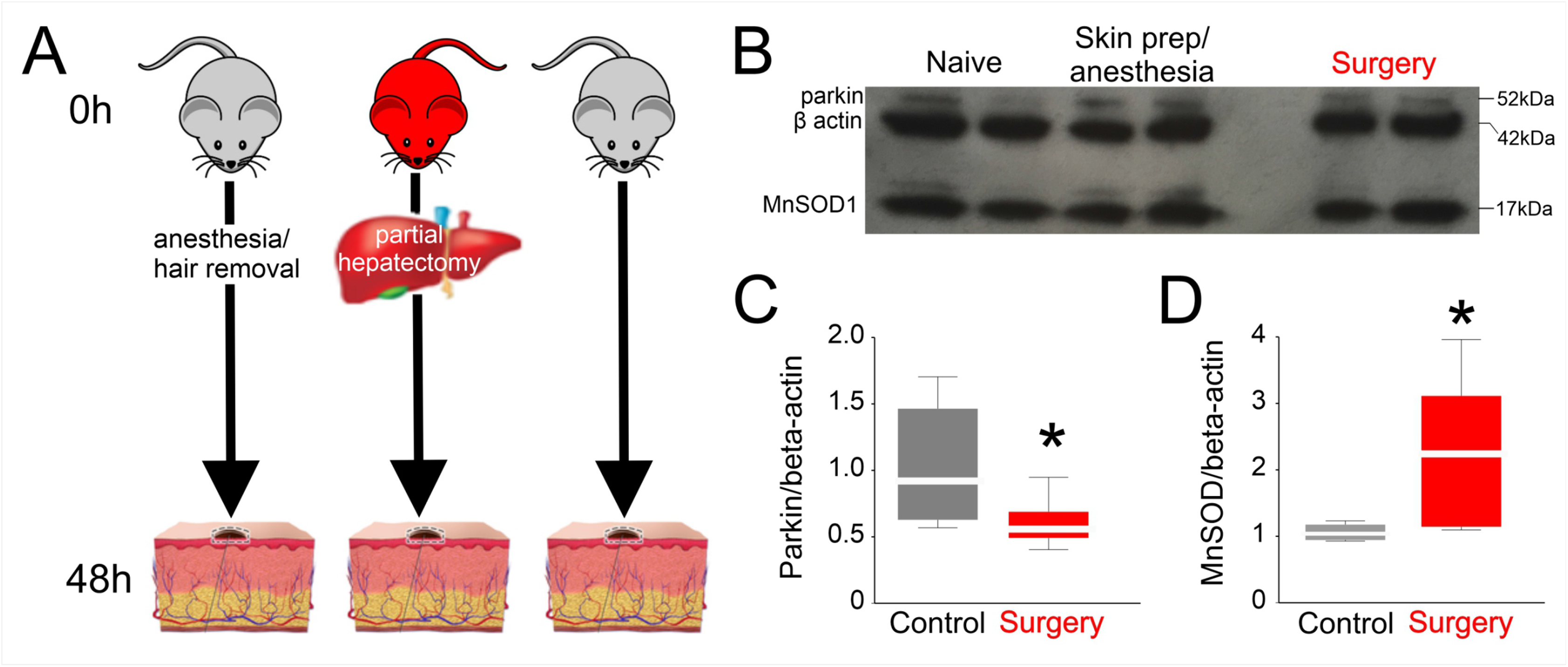
Postoperative differences in protein expression related to oxygen delivery intervention. A. Proteins that were differentially expressed after surgery in non- DO_2_ achievers and DO_2_ achievers (median confidence score:63.7 (27.1-128.5)). Heatmap data is shown for relative changes for within protein comparisons, combining data for achievers versus non-achievers. B. Summary of potential molecular interactions using Ingenuity Pathway Analysis, which were assessed by comparing protein expression levels before and after surgery for nonDO_2_ achievers and DO_2_ achievers. Links are color-coded as orange: leads to activation, blue: leads to inhibition, gold: findings inconsistent with state of downstream protein, grey: effect not predicted. Proteins that are experimentally validated are indicated by a black arrow. The orange color of the target upstream regulator and biological function shown in the center implies ‘‘activation,’’ and the shade of the color implies confidence in activation with darker shades implying more confidence in the prediction. C. Proteins involved in smooth muscle migration were upregulated in patients in whom oxygen delivery target was achieved. D. Leukocyte activation pathways were upregulated in patients in whom oxygen delivery target was achieved. E. In patients who failed to achieve their preoperative DO_2_, epithelial necrosis was identified through relative over-expression of proteins involved in oxidative stress (superoxide dismutase, PARK7).

## Discussion

Our state-of-the-art proteomic analysis of serial skin biopsies revealed reproducible molecular and cellular signatures of end-organ injury after major surgery. Proteomic changes related to perioperative oxygen delivery revealed a link between lower oxygen delivery, oxidative stress and reparative processes previously identified by gene knockout experiments. The proteomic data from patients was verified by immunohistochemistry using selected validated antibodies and further supported by immunoblots of abdominal wall skin samples obtained from a translational murine model of major surgery. Taken together, these data suggest that serial skin biopsies provide mechanistic insight into organ injury after surgery.

Despite skin being the largest organ in the body and also being readily accessible, human proteomic studies are scarce.^28^ In part, this reflects that skin has presented significant challenges for conventional proteomic techniques due to its high lipid content, insolubility and extensive cross-linking of proteins, all of which complicate the isolation and digestion of proteins for analysis using mass spectrometry techniques.^16^ This study was facilitated by our recent development of on-line fractionation and optimised acquisition protocols utilising ion mobility separation technology has significantly increased the scope for protein identification ten-fold from a single punch biopsy.^16^

A landmark surgical study that employed SOMAscan proteomic technology identified 564 proteins in blood plasma from older adults undergoing elective surgery whose expression levels were consistently changed after surgery. ^29^ Our study extends that work by focusing on skin, an end-organ that is vulnerable to reduced oxygen delivery, a key driver of postoperative complications. We demonstrate that tracking proteomic changes using skin biopsies is acceptable to surgical patients, easy to perform and provides integrated mechanistic insights into end-organ physiology that is seldom reflected by changes in circulating proteins in blood.

The proteomic profiles revealed by our study have captured components from each of the four dynamic overlapping processes required for the coordination of successful healing of skin wounds.^30^ Briefly, haemostasis is triggered once platelets encounter collagen and the extracellular matrix. The development of fibrin clot and release of clotting factors, growth factors and cytokines initiates the inflammatory phase, which lasts for ~48h (i.e. the timeframe over which the two skin punch biopsies were taken in our study). The inflammatory phase is heralded by the chemotaxis of neutrophils and monocyte/macrophages to commence phagocytosis of cellular debris and pathogens, assisted by the activation of mast cells and fibroblasts. New extracellular matrix is then created by deposition of collagen by fibroblasts and further remodelling facilitated by TGFβ, proteoglycans, fibronectin and protease inhibitors.^31^ Our proteomic profiling ceased before the process of proliferation and epithelisation is widely held to be completed, which requires angiogenesis and neovascularisation.

Canonical pathway analysis highlighted that acute phase proteins and sirtuin signaling were markedly upregulated in patients who failed to maintain preoperative oxygen delivery, suggesting that reduced oxygen delivery results in excess skin inflammation and an exaggerated stress response. The sirtuins are a family of NAD+ dependent comprising class III histone deacetylases regulating the metabolic and transcriptomic response to oxidative stress in skin.^21^ Activation of sirtuins 1-3 accelerate wound healing.^32^ At the individual protein level, we found a coordinated upregulation of antioxidant proteins including mitochondrial heat shock protein, deglycase DJ*-*1 and the mitochondrial antioxidant manganese superoxide dismutase. These changes were exclusively observed in patients where preoperative oxygen delivery was not achieved within 8h of completing surgery, suggesting strongly that failure to supply skin with adequate oxygen delivery during the early perioperative period results in delayed or persistent cellular metabolic stress for at least 36h after surgery. Our proteomic and immunohistochemical analysis also found, across the entire population, decreased postoperative expression of CD44, the epidermal expression of which is required for optimal wound healing by maintaining epidermal elasticity and resistance to stretch, as well as keratinocyte proliferation and differentiation.^33^

Very few studies have focused on the molecular mechanisms underpinning the association between adequate perioperative oxygen delivery and reduced morbidity and/or mortality. Stroke volume guided fluid and low dose inotropic therapy was associated with improved oxygen delivery, microvascular flow and tissue oxygenation but had no impact on global (plasma) measures of inflammation after major surgery.^34^ By contrast, proteomic analysis of skin biopsies obtained after surgery from patients who achieved their preoperative oxygen delivery target demonstrated a coordinated leukocyte activation response that was absent in non-achievers. The ubiquitin–proteasome system degrades intracellular proteins into peptide fragments that can be presented by major histocompatibility complex (MHC) class I molecules. Unique to the non-achiever group was the increased expression of PSMB9, one of two critical immunoproteasome subunits that degrade cell proteins to generate peptides for antigen presentation exclusively during acute inflammation.^35^ Immunoproteasome expression is therefore closely associated with T-cell infiltration, through cytokine production and T cell expansion and/or survival, by regulating apoptotic machinery and/or transcription factor activation.^36^ Achievement of preoperative oxygen delivery was also associated with increased expression of the antimicrobial peptidoglycan recognition protein-1 (PGLYRP-1), which is expressed primarily in the granules of polymorphonuclear leucocytes and is present in neutrophil extracellular traps.^37^ PGLYRP-1 is directly bactericidal for both Gram-positive and negative bacteria by interacting with cell wall peptidoglycan, rather than permeabilizing bacterial membranes.^38^ The observed increase in expression of kininogen, through which kinins are formed in the skin by the enzymatic action of tissue kallikrein, is consistent with its’ role in skin homeostasis, wound healing and organ dysfunction more widely.^39^

We also noted that increased expression of thrombospondin-1 and plasminogen in both groups. The presence of thrombospondin-1 (TSP-1), a potent chemotactic factor for monocytes and neutrophils, in skin biopsy samples 48h after surgery is consistent with its role in the early inflammatory phase of wounds; absence of TSP-1 delays wound healing.^40^ However, given that we did not acquire further biopsy samples beyond 48h of surgery, we cannot exclude that the deleterious effects of persistently elevated expression of wound bed TSP1 prevents effective wound healing over the longer term.^41^ The early presence of plasminogen in postoperative biopsy samples from both groups is also consistent with the critical role of this potent serine protease in triggering the resolution of inflammation and activating the proliferation phase of wound repair.^42^

A strength of our study design was the within-subject comparison of punch biopsies within a similar abdominal area, thereby serving as a robust internal control. The masked analysis of samples to trial allocation is another strength. There are several limitations of this ‘first-in-man’ study of surgical patients. Samples obtained as part of a randomised controlled study in a specific subset of patients at higher risk of perioperative morbidity may not be generalizable across all surgical populations. The proteins selected for immunohistochemical validation represent only a small subset of the proteins impacted by surgery and were chosen based on the availability of validated antibodies for immunohistochemistry, systems biology analysis, and literature supporting a link with skin-specific pathology. Due to cost and patient-related constraints, further samples beyond 48h after surgery were not obtained during the proliferative reparative phase. Usually, punch biopsies heal 5-10 days after the procedure, which would have lent itself to monitoring the rate of healing if the patients had been guaranteed to reside in hospital until the punch biopsy site had healed. A derivation-validation approach correlated with postoperative outcomes would add further value.

## Conclusion

Serial skin biopsies identified novel surgery-specific protein changes in adults undergoing major elective, noncardiac surgery. Selected proteins validated in a larger cohort correlated with established physiologic precursors for morbidity and mortality. This approach may identify potential biomarkers with clinical utility, particularly as we observed upregulation of biomarkers over time. Proteomic analysis of skin biopsies provides in-depth understanding of perioperative end-organ dysfunction, including the potential development of precision medicine-guided treatments for wound healing after major surgery.^43^

## Data Availability

Data requests to GL Ackland

## Acknowledgments

This work is supported by the UK NIHR Great Ormond Street Hopsital Biomedical Research Centre. The views expressed are those of the author(s) and not necessarily those of the NHS, the NIHR or the UK Department of Health.

